# Evaluating specialist intensive support teams for adults with intellectual disabilities who display behaviours that challenge: The IST-ID mixed methods study

**DOI:** 10.1101/2022.05.16.22275150

**Authors:** Angela Hassiotis, Athanasia Kouroupa, Leila Hamza, Louise Marston, Renee Romeo, Nahel Yaziji, Ian Hall, Peter E Langdon, Ken Courtenay, Laurence Taggart, Nicola Morant, Vicky Crossey, Brynmor Lloyd-Evans

## Abstract

**Background:** Intensive Support Teams (ISTs) are recommended for individuals with intellectual disabilities who display behaviours that challenge. However, there is currently little evidence about the clinical and cost effectiveness of IST models operating in England.

**Aims:** To investigate the clinical and cost effectiveness of IST models.

**Methods:** We carried out a cohort study to evaluate the clinical and cost-effectiveness of two previously identified IST models (independent and enhanced) in England. Adult participants (n=226) from 21 ISTs (10 independent and 11 enhanced) were enrolled. The primary outcome was change in challenging behaviour between baseline and 9 months measured by the Aberrant Behaviour Checklist-Community 2.

**Results:** We found no statistically significant differences between models for the primary outcome (adjusted β: 4.27; 95% CI: -6.34 to 14.87; p=0.430) or any secondary outcomes. Quality Adjusted Life Years (0.0158; 95% CI: -0.0088 to 0.0508) and costs (£3409.95; 95% CI: -£9957.92 to £4039.89) of the two models were comparable.

**Conclusions:** The study provides evidence that both models were associated with clinical improvement for similar costs at follow-up. We recommend that the choice of service model should rest with local services. Further research should investigate the critical components of IST care to inform the development of fidelity criteria, and policy makers should consider whether roll out of such teams should be mandated.

**Study registration number:** ClinicalTrials.gov NCT03586375; IRAS 239820; National Institute for Health Research (NIHR) Central Portfolio Management System (CPMS) 38554.

## Introduction

Approximately 18% of adults with intellectual disabilities (lifelong limitations in adaptive functioning evident in early life) living in the community display aggression, self-injury, property destruction or other socially inappropriate behaviours in their lifetime(1,2). Some 24,000 adults with intellectual disabilities are at risk of being admitted to specialist psychiatric Assessment and Treatment units often due to the display of such behaviours(2,3).

Research suggests that these individuals are subject to unnecessary long-term psychotropic medication use, poorer health, abuse and social exclusion(1,4). International studies indicate that adults with intellectual disabilities are more likely to visit the emergency department for psychiatric issues(5), return to the emergency department within 30 days of discharge(6), be in long-term inpatient care and experience premature mortality(7). Failure to effectively address behaviours that challenge before a crisis arises causes significant distress and burden to families and consequent breakdown of placements(8,9), in addition to significant healthcare and societal costs. A recent census of the Transforming Care Programme(10) in England, a national initiative to drive improvements in the care of people with intellectual disabilities who display behaviours that challenge, indicated minimal change in relation to the number of inpatient admissions, length of hospital stay, out-of-area placements and antipsychotic medication use, confirming concerns about the lack of progress in the care of this population group across the country(11). Intensive Support Teams (ISTs) are community services that complement the community intellectual disability services and have been in operation since the early days of community care(12,13). However, there is little evidence to recommend a preferred IST model, and there are no nationally specified outcomes for IST care. The National Institute for Health and Care Excellence(14) recognised the importance of such specialist treatment services but did not find sufficient evidence that they were clinically effective or that they reduced costs. Hassiotis et al. have reported the typology of ISTs which led to the identification of two models, independent and enhanced(15). The aim of the present study was to examine the clinical and cost effectiveness of the two IST models at 9 months follow up.

## Method

### Study design

The primary and secondary outcomes were collected at baseline and 9 months follow up. At the time of study completion, there were UK-wide public health measures implemented due to the covid-19 pandemic and in person assessments could not take place between March 2020 and January 2021.

### Service and participant recruitment

The research team prepared a matrix of all identified IST services in England stratified by model type, caseload size and area. The service managers of ISTs representing the two models, were randomly invited to take part in the study. If refused or not responded, the next service in the matrix was approached until the required number of ISTs and participants were enrolled. The inclusion criteria of the study for services were: 1. ISTs operational for at least one year; 2. ISTs funded for the duration of the study; and for service user participants these were: 3. Adults aged 18 years or over with a clinical diagnosis of mild to profound intellectual disabilities; 4. being under the care of an IST (either model) including new referrals. Those with a primary diagnosis of personality disorder or substance misuse, or a clinical decision that a taking part in the study would be inappropriate due to risks were excluded. Potential participants and their family/paid carers were approached by researchers and, where available, staff from the Clinical Research Networks to seek expressions of interest to take part in the study.

### Consent statement

Participants provided written and/or audio recorded verbal consent for in person or online assessments, respectively. For participants with intellectual disabilities who did not have capacity to make an informed decision about taking part in this study, we obtained written and/or audio recorded agreement from a personal/nominated consultee.

### Ethics Statement

This study was performed in accordance with the Declaration of Helsinki. Ethical approval was provided by the London Bromley Research Ethics Committee (18/LO/0890).

### Outcomes

The primary outcome was change in challenging behaviour as measured by the carer reported Aberrant Behaviour Checklist-Community version 2 (ABC-C)(16). Secondary outcomes were mental health comorbidity (PAS-ADD Checklist)(17), clinical risk (Threshold Assessment Grid; TAG)(18) and quality of life (QoL-Q)(19). The ABC-C, PAS-ADD and QoL-Q have been validated for use with people with intellectual disabilities. The TAG is widely used in clinical practice to capture clinical risk in mentally ill patients and has been used previously in a population with intellectual disabilities(20). Quality-adjusted life years (QALY) were derived from the EQ-5D-5L(21) scores. If the participant with intellectual disability had sufficient reading ability and aided by the researcher, a self-report version of the EQ-5D-5L was administered. It is recommended that the proxy EQ-5D-5L should also be completed for adults with intellectual disabilities(21). Use of hospital and community services was using the study adapted Client Service Receipt Inventory (CSRI)(22) covering the previous 6 months. At 9 months’ follow-up, service use for the previous 6 months was extrapolated to 9 months.

### Additional information

We also collected sociodemographic details, the Adaptive Behaviour Scale – short-form (SABS)(23) as proxy of intellectual disability (higher scores indicate mild intellectual disability), medication use, number of hospital admissions and changes in accommodation.

### Sample size

The sample size was calculated to detect a difference of 0.45 standard deviation (SD) in primary outcome score. Assuming two IST models, this required 96 participants per group (192 in total) with 5% significance (two-sided) and 80% power and an intraclass correlation coefficient of 0.02(24). After inflation for 15% loss to follow-up, the estimated sample size was 113 participants per model (226 participants in total).

### Statistical analyses

A detailed statistical plan was developed a priori and reviewed by the oversight Study Steering Committee. All analyses were carried out using Stata/IC v16.0(25). All hypothesis testing was conducted using a two-sided significance level of 5%, with corresponding 95% confidence intervals.

#### Clinical effectiveness

The primary outcome was estimated using a mixed effects linear regression model with change in ABC-C score as the outcome, a fixed effect for IST type as the main exposure, and a random effect for IST to account for clustering within services. We carried out unadjusted modelling, then age, gender, accommodation type, level of ID (SABS score), level of risk (baseline TAG), presence of autism and/ or Attention Deficit Hyperactivity Disorder (ADHD), number of physical comorbidities, PAS-ADD organic, affective, and psychotic positive were identified as potential confounders and were included in an adjusted model. Continuous secondary outcomes were analysed using statistical models analogous to those for the primary outcome. Binary outcomes were analysed using mixed effects logistic regression models and were unadjusted. Analyses of secondary outcomes were considered exploratory. Predictors of missingness of the primary outcome were examined using mixed effects logistic regression. Where there were up to 20% missing items for the ABC-C, TAG and QOL-Q, they were replaced by the mean score of items present. Where items were missing for the PAS-ADD, they were replaced by a code indicating the participant was negative for the given condition.

### Health economic analysis

A detailed health economic analysis plan was also developed a priori and reviewed by the oversight Study Steering Committee and followed similar principles to the statistical analysis plan regarding assumptions. All analyses were carried out using Stata/IC v16.0(25).

#### Economic evaluation

##### Perspective

The cost effectiveness analysis adopted the perspectives of health and social care which cover hospital and community health, social care services, and included voluntary support from outside the family funded by social services and provided by not-for-profit organisations. Wider societal perspective also includes the cost of unpaid support to the participant by family and friends.

##### Valuation of resource use

Costs of the IST service models were derived by combining data on annual salary, working time, overheads, number of sessions with participants, information on caseloads and referrals over 12 months. Travel costs to home visits were included where this was noted. The annual cost was then weighted to derive a cost per study participant for each IST model over 9 months. Unpaid support costs were calculated using the market price approach, the hourly rate of a home care worker was used for those not in employment and if employed, the carer hourly wage rate. All unit costs were for the financial year 2020/2021.

##### Cost-effectiveness

We analysed differences in mean health and social care costs and wider societal costs at 9 months in turn between the IST models by regressing total cost from each perspective on IST model, baseline costs, total ABC-C score, health-related quality of life tariffs, and a range of clinical and socio demographic indicators. Non-parametric bootstrapping was used to estimate 95% confidence intervals for mean costs. Significance was set at p<0.05.

Cost-effectiveness was explored using the net benefit approach(26,27) with effectiveness measured in terms of the primary outcome measure (ABC-C score) and QALY gains were derived by developing value sets from the EQ-5D-5L by means of a crosswalk to the EQ-5D-3L value sets(28) at each time point. Uncertainty around the cost and effectiveness estimates was represented by cost-effectiveness acceptability curves(29).

In sensitivity analyses we examined whether adjustment for baseline characteristics found to be associated with missing data affected the main findings. Those variables identified as significantly associated with missingness were then added to the baseline covariates used in main analyses and new ICERs were re-estimated.

### Covid-19 impact and adaptations

Three NHS sites withdrew their participation when the NIHR suspended all non-Covid-19 related research in March 2020. In order to carry on with recruitment, we applied for and received ethical approval to complete the consent process and research assessments remotely using digital platforms (e.g., Zoom, phone calls, scanned copies via email). Challenges to the study included digital poverty (e.g., lack of computer/smart phone); insufficient knowledge of using digital platforms and where a participant could receive support from if doing so; difficulty in assessing whether a service user with intellectual disabilities had sufficient verbal ability to provide consent remotely, and delays in obtaining contact details for consultees.

## Results

### Clinical outcomes

The STROBE diagram (Figure 1) presents the participant flow into the study. Enrolment took place between August 2018 and May 2020 with last participant assessment in January 2021. There was an 8% attrition rate due to the following reasons: 1. uncontactable (n=11); 2. death (n=2 of which one was due to Covid-19); 3. missing follow-up assessment window (n=2); 4. hospitalisation (n=1); 5. imprisonment (n=1); and 6. excessive stress during the pandemic (n=1).

**Figure 1.**
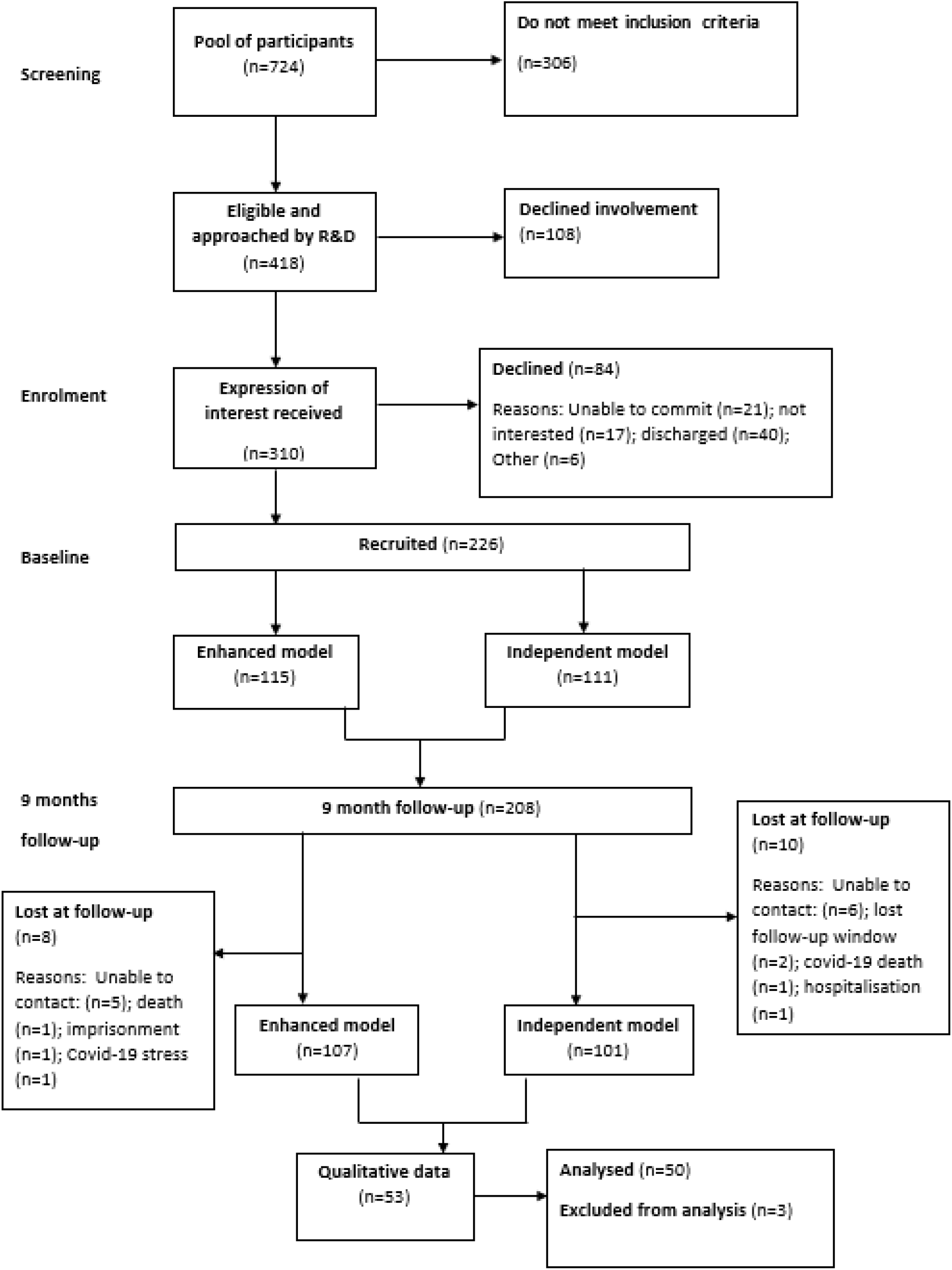
STROBE Diagram.

Demographic characteristics of adults with intellectual disabilities per IST model at baseline and 9-month follow-up are presented in Table 1. The median age of participants was 29 years old (IQR 23, 39) and the majority were single male of white ethnicity. More than 60% of participants had comorbid developmental disorders. The whole cohort level of adaptive ability was 52 (SD=24). Participants in the two models differed in the number of reported sensory problems (enhanced 52% vs. independent 68%, p=0.018) and education status (enhanced 45% vs. independent 32%, p=0.035). At follow up, participants were more likely to be receiving care from the enhanced IST compared to those still in contact with the independent IST (enhanced n=78; 74% vs. independent n=45; 45%, p-value=0.001). The median time adults with intellectual disabilities were seen from the enhanced ISTs was 20 months [IQR (12, 33)] compared to 13 months in independent ISTs [IQR (10, 22)].

**Table 1.** Baseline sociodemographic and clinical characteristics by IST model

|  | **Baseline** |  |  |
| --- | --- | --- | --- |
|  | **Enhanced (n=115)**<br>**Mean (SD) or N (%)** | **Independent (n=111)**<br>**Mean (SD) or N (%)** | **Total (n=226) Mean**<br>**(SD) or N (%)** |
| Age (years) median (IQR) | 30 (24, 41) | 28 (22, 38) | 29 (23, 39) |
| Age 25+ | 81 (70) | 70 (63) | 151 (67) |
| Male | 80 (70) | 75 (68) | 155 (69) |
| **Ethnicity** |  |  |  |
| White | 90 (78) | 91 (82) | 181 (80) |
| Black | 11 (10) | 12 (11) | 23 (10) |
| Asian | 12 (10) | 5 (5) | 17 (8) |
| Other | 2 (2) | 3 (3) | 5 (2) |
| **Short Adaptive Behaviour Scale** | 50 (25) | 56 (22) | 52 (24) |
| **Neurodevelopmental disorder** |  |  |  |
| ASD<sup>1</sup> or ADHD<sup>2</sup> | 72 (63) | 72 (65) | 144 (64) |
| Neither ASD or ADHD | 33 (29) | 33 (30) | 66 (29) |
| Both ASD and ADHD | 10 (9) | 6 (5) | 16 (7) |
| **Aetiology of intellectual disabilities** |  |  |  |
| Unknown | 95 (83) | 86 (77) | 181 (80) |
| Other | 12 (11) | 20 (18) | 32 (14) |
| Down's or Fragile X Syndrome | 7 (6) | 5 (5) | 12 (5) |
| **Physical health problems** |  |  |  |
| Mobility problems | 45 (39) | 40 (36) | 85 (38) |
| Sensory problems* | 60 (52) | 75 (68) | 135 (60) |
| Epilepsy | 33 (29) | 25 (23) | 58 (26) |
| Incontinence | 41 (36) | 35 (32) | 76 (34) |
| Other physical health | 33 (29) | 35 (32) | 68 (30) |

|  | Baseline |  |  |
| --- | --- | --- | --- |
|  | Enhanced (n=115)<br>Mean (SD) or N (%) | Independent (n=111)<br>Mean (SD) or N (%) | Total (n=226) Mean<br>(SD) or N (%) |
| problem(s) |  |  |  |
| **Marital status** |  |  |  |
| Single | 113 (98) | 111 (100) | 224 (99) |
| Married | 2 (2) | - | 2 (1) |
| Divorced | - | - |  |
| **Living situation** |  |  |  |
| Alone or with partner<br>+/- children | 23 (20) | 22 (20) | 45 (20) |
| With parents/relatives | 34 (30) | 24 (22) | 58 (26) |
| Other (e.g., sheltered<br>accommodation,<br>group living, hospital) | 58 (50) | 65 (59) | 123 (54) |
| **Accommodation** |  |  |  |
| Time in current<br>address (months)<br>median (IQR) | 48 (12, 144) | 36 (11, 146) | 47 (12, 144) |
| Family home | 38 (33) | 30 (27) | 68 (30) |
| Supported living | 34 (30) | 47 (42) | 81 (36) |
| Residential | 33 (29) | 27 (24) | 60 (27) |
| Independent | 9 (8) | 7 (6) | 16 (7) |
| Less than 6 months in<br>current<br>accommodation | 13 (11) | 19 (17) | 32 (14) |
| **Main source of**<br>**income** |  |  |  |
| Salary/ wage | - | 1 (1) | 1 (0.4) |
| Family support | 35 (30) | 27 (24) | 62 (27) |
| State benefits | 115 (100) | 111 (100) | 226 (100) |
| **Occupational and**<br>**activities status** |  |  |  |
| None | 67 (58) | 73 (66) | 140 (62) |
| Full/ part-time<br>employment | 2 (2) | 2 (2) | 4 (2) |
| Education, day<br>centre, looking after<br>home, other* | 52 (45) | 35 (32) | 87 (38) |
| Voluntary work | 6 (5) | 6 (5) | 12 (5) |

<sup>1</sup>ASD: Autism Spectrum Disorder, <sup>2</sup>ADHD: Attention deficit hyperactivity disorder \*: p-value <0.05

### Primary outcome

Baseline mean total ABC-C scores were similar between IST models [enhanced 64 (SD=34), independent 62 (SD=32)] (Supplementary Table 1). The mean ABC-C scores were lower at 9 months for both IST models: enhanced 56 (SD=34), independent 49 (SD=32) (Supplementary Table 1). Both unadjusted and adjusted analyses found no statistically significant difference in total ABC-C score change between IST models at 9 months (adjusted β: 4.27; 95% CI: -6.34, 14.87) (Table 2). The only predictors of missingness were physical health conditions.

**Table 2.** Change in clinical outcomes of ISTs at 9 months in terms of independent IST

|  | Difference between IST model |  |  |  |
| --- | --- | --- | --- | --- |
|  | Unadjusted |  | Adjusted |  |
| Outcome | Estimate | 95% CI | Estimate | 95% CI |
| **Primary outcome** |  |  |  |  |
| Change in ABC-C total ($\beta$) | 3.08 | -7.32, 13.48 | 4.27 | -6.34, 14.87 |
| Adjusted for time between baseline and 9-month data collection ($\beta$) | 2.75 | -7.56, 13.06 | 3.62 | -6.99, 14.22 |
| Change in ABC-C Irritability ($\beta$) | 1.14 | -2.41, 4.69 | 2.09 | -1.67, 5.86 |
| Change in ABC-C Lethargy ($\beta$) | 1.40 | -1.77, 4.58 | 1.31 | -1.82, 4.44 |
| Change in ABC-C Stereotypic behaviour ($\beta$) | -0.05 | -1.44, 1.34 | 0.69 | -0.77, 2.14 |
| Change in ABC-C Hyperactivity ($\beta$) | -0.07 | -3.54, 3.39 | -0.11 | -3.68, 3.45 |
| Change in ABC-C Inappropriate speech ($\beta$) | 0.83 | -0.10, 1.77 | 0.37 | -0.86, 1.60 |
| **Secondary outcomes (unadjusted)** |  |  |  |  |
| PAS-ADD Organic condition (OR) | 1.09 | 0.39, 3.02 |  |  |
| PAS-ADD Affective or neurotic disorder (OR) | 0.91 | 0.32, 2.59 |  |  |
| PAS-ADD Psychotic disorder (OR) | 1.08 | 0.21, 5.50 |  |  |
| Change in TAG ($\beta$) | 1.11 | -0.35, 2.57 | 1.12 | -0.44, 2.68 |
| Change in QOL-Q ($\beta$) | -0.75 | -3.62, 2.11 | -2.63 | -5.65, 0.40 |

### Secondary outcomes

No statistically significant differences were found in any of the secondary outcomes between IST models at 9 months (Table 2 and Supplementary Table 1).

### Medication use

The mean number of medications prescribed at baseline was the same for both models (n=5). At follow-up, the mean number was slightly reduced in the independent model (4 vs. 5 in the enhanced). At baseline, psychotropic medication was prescribed at similar proportions in both models [enhanced and independent respectively: antipsychotic (17% vs. 18%), other psychotropic (35% vs. 30%)]. The relative proportions of prescribed antipsychotics did not change at follow up [enhanced and independent respectively: antipsychotic (18% vs. 20%), other psychotropic (31% vs. 32%)].

### Psychiatric hospitalisations and change in accommodation

Over the study duration, 8 participants in the enhanced model and 11 participants in the independent were admitted to a psychiatric unit as a result of a mental health crisis. Nine (20%) participants moved accommodation during the study period.

### Cost evaluation

The average annual cost of teams in the enhanced model, was £612,612 (£4,980 per case) whilst the average annual cost of a team in the independent model was £647,812 (£10,122 per case) (Supplementary Table 2).

#### Service use

From an NHS/Personal Social Services perspective, the mean total cost over 9 months was £22915.6 for the independent model and £19037.6 for the enhanced; the adjusted mean difference in costs was not statistically significant (£446.55, 95% CI -£(−5637.60 to £7519.30).

From a societal perspective the mean total cost over 9 months was £31850.8 for the independent model and £29852.8 for the enhanced. The adjusted mean difference in costs was not statistically significant (−£855.8095% CI -£8342.54 to £6059.69).

The mean use of inpatient, outpatient, and day-patient health services over the 9-month follow-up period are reported in Supplementary Table 3. Duration of inpatient stay, outpatient attendances, day hospital contacts and emergency (A&E) attendance, were broadly similar for both models. Notably, participants in the independent model spent longer on average, as inpatients than those in the enhanced model [8.63 (SD 39.98) vs. 5.26 (SD 28.10), respectively]. Participants in the independent model had on average more contacts with their general practitioner than participants in the enhanced model (mean (SD) 4.88 (14.20) vs. (SD) 3.47 (3.96) attendances respectively) though these were not statistically significant.

#### Cost-effectiveness

There are no statistically significant differences in QALYS in any of the comparisons of the service models at 9 months (Supplementary Table 4). Results from the regression analysis using the two outcomes of total ABC-C score and QALYs are summarised, and the incremental cost effectiveness ratios (ICERs) reported in Supplementary Table 5.

Probability estimates were plotted for a range of implicit monetary values attached to improvements in total ABC-C score and QALY gain over 9 months under an NHS and societal perspective in turn (Supplementary figure 1 - 4).

The independent model had a low likelihood (∼ 50%) of being more cost-effective than the enhanced model if decision makers were not willing to pay anything for a unit improvement in the total ABC-C score. The likelihood of cost-effectiveness rose to 60% if a decision maker was willing to pay £500 for a unit improvement in total ABC-C score; and remained at 70% if willingness to pay for an improvement in total ABC-C score rose to £1000. Under a broader perspective which includes cost of unpaid support by family and friends, the probability of the independent model being cost-effective, compared with the enhanced at the standard NICE-preferred willingness-to-pay levels of £20,000–30,000 per QALY, was 52%. It is therefore unlikely that there are any economic gains from choosing one model of care over another. Controlling for factors contributing to missing data in health and social care costs in sensitivity analyses, did not alter the findings of the main analyses.

## Discussion

The study showed that both IST models currently in operation in England were associated with reduction in behaviours that challenge at 9 months follow-up with comparable costs. Most recently, Dodd, Laute and Daniel(30) reported on a new model of integrating intensive support service with an inpatient unit for adults with complex needs including behaviours that challenge, at risk of admission. The new remodelled service received a significant number of referrals and appears to have had an increasing efficacy in preventing admissions from two thirds in 2016 to 90% in 2020. Placement of patients was maintained in up to 80% of cases. However, although duration of inpatient care was reduced from 18 months to less than 12 months as a result of the intensive support, delayed discharges remained mostly as a consequence of lack of suitable accommodation. Bohen and Woodrow(31) and Mottershead and Woodrow(32) published initial findings for the Dynamic Support Database clinical support tool. Such work, though in its early stages, is promising in not only the identification of patients likely to require admission, but also pointing towards prevention and intervention strategies that could contribute to the refinement of the specification of ISTs. Iemmi et al(33) reported on the economic evaluation of one IST delivering positive behaviour support to five patients. Their study claimed that the IST contributed to improved outcomes in challenging behaviour and increased community participation at a total cost of health and social care services of £2,296 per week. Hassiotis et al(34) highlighted that a specialist behaviour team was cost neutral when service use was taken into consideration.

The health economic evaluation of this multicentre study showed that the service costs of enhanced ISTs are not significantly different the independent ISTs, and neither are the health-related quality of life gains. However, as the financial burden of behaviours that challenge remains substantial(35,10,14), it is likely that costs could be offset by clinical improvements associated with IST care. Cost per case would range between £4,980 and £10,122 for the enhanced and independent model respectively which is significantly cheaper than current costs within the NICE cost acceptability threshold. However, the magnitude of that change would have to exceed the unit improvement in the total ABC-C in order to be considered as clinically significant. Previous research indicates that experts by experience expect larger differences as meaningful than those reported in existing publications(36).

### Strengths and limitations

This study is the first, to our knowledge, to have systematically evaluated IST models in England. It was fully powered with very good retention of participants (less than 10% attrition) the Covid-19 pandemic notwithstanding. ISTs were representative of such services in England and the study participants representative of the population on IST caseloads, which minimise the risk of bias. The findings from this study are highly relevant to the support of very vulnerable individuals with intellectual disabilities in the community and potentially applicable to other UK countries where they seek to establish similar approaches to the acute or preventive management of behaviours that challenge.

The study also has limitations. Firstly, responses might be subject to respondent social desirability bias. Second, this was not a randomised controlled trial so there may have been differences between groups which we were unable to measure and adjust for in the analyses. Third, the turnover of paid carers may have affected the reporting of behaviours that challenge if the carer had not known the person with intellectual disabilities for long enough. Fourth, the lack of statistical significance in clinical outcomes between models may be an indication that adults who are referred during a crisis will recover substantially in the short to medium term (regression to the mean) especially where the disorder is remitting/relapsing. Fifth, as we did not recruit participants at the point of referral to the IST we must be cautious about the change that was achieved as it has not taken into account any improvements made prior to study entry. Sixth, there may have been some impact due to the Covid-19 pandemic as 131 follow-up interviews were conducted from March 2020 to January 2021, but we were unable to fully adjust for it. For example, it may have exacerbated behaviours that challenge or affected the patterns and intensity of service use in both models. This is especially important, given the current disproportionate impact of Covid-19 on people with intellectual disabilities, including higher death rates(37).

## Conclusion

Whilst there is currently a policy view that ISTs may be more effective than CIDS in averting adverse outcomes of behavioural crises and the clinical improvements shown in the study are promising, further high-quality evaluation of IST versus CIDS is needed to confirm this. We also need to investigate the critical ingredients of effective IST care and understand how best ISTs may work with and fit into the wider mental health service system. A potential starting point would be a clear delineation of IST role in a crisis pathway to include protocols for rapid response with innovative and flexible approaches to short-term assessment and treatment. From a public health and policy perspective, whilst commissioners can choose which IST model is relevant to their localities, any service developments must to be rooted in evidence and be able to accommodate a host of solutions from care delivery (remote or in person), expert by experience input and crisis prevention in order to enhance good practice and more importantly improve the patient experience of care.

## Supporting information

Supplement to main document 5 tables and 4 figures

## Data Availability

Anonymised data included in the survey are available from the authors on request and subject to internal review of proposals.

## Acknowledgements

The authors are grateful to the study participants who participated in the study. The research team is also grateful to the NIHR Clinical Research Network (CRN), all 21 Research and Development offices, and to the Project Advisory Group of experts by experience for their support and commitment to the study. In particular, we acknowledge the support and contribution of Narender Kaur, Lorna Bryan, Janet Seamer, Sandy Smith and Brendan Leahy from the Camden Disability Action (Synergy). We would also like to thank the Study Steering Committee, chaired by Professor Nora Grace, for their oversight of the study. Finally, we are grateful to previous members of the research team: Amy Walsh, Isobel Harrison, Dr Victoria Ratti, Jessica Budgett, Peiyao Tang for their contribution to delivering the IST-ID study.

## Author contribution

A.H. is the chief investigator of this study. A.K and L.H prepared the manuscript with A.H. A.K and L.H. carried out data collection. L.M. performed the analysis of clinical data while R.R. and N.Y. performed analyses of the health economic data. All authors have contributed to all iterations of the manuscript and its intellectual content.

