## Supplement to main document 5 tables and 4 figures for "Evaluating specialist intensive support teams for adults with intellectual disabilities who display behaviours that challenge: The IST-ID mixed methods study"

### Supplementary Material

#### Tables

**Supplementary Table 1** Primary and secondary outcome scores at baseline and 9 months

|  | Baseline | | 9 months | |
| --- | --- | --- | --- | --- |
| Outcome | **Enhanced (n=115)**  **Mean (SD) or N (%)** | **Independent (n=111) Mean (SD) or N (%)** | **Enhanced (n=107)**  **Mean (SD) or N (%)** | **Independent (n=101) Mean (SD) or N (%)** |
| ABC-C^1^ Total | 64 (34) | 62 (32) | 56 (34) | 49 (32) |
| ABC-C Irritability | 21 (11) | 20 (11) | 17 (11) | 15 (10) |
| ABC-C Lethargy | 14 (9) | 14 (10) | 12 (9) | 11 (9) |
| ABC-C Stereotypic behaviour | 6 (6) | 5 (5) | 6 (5) | 5 (5) |
| ABC-C Hyperactivity | 19 (13) | 18 (11) | 17 (11) | 14 (11) |
| ABC-C Inappropriate speech | 4 (4) | 5 (4) | 4 (4) | 4 (4) |
| PAS-ADD^2^ | | | | |
| Organic condition | 14 (12) | 16 (14) | 8 (8) | 8 (8) |
| Affective or neurotic disorder | 22 (19) | 26 (23) | 12 (11) | 10 (10) |
| Psychotic disorder | 4 (3) | 11 (10) | 3 (3) | 3 (3) |
| TAG^3^ | 14 (5) | 13 (5) | 14 (5) | 12 (5) |
| QOL-Q^4^ | 70 (11) | 69 (8) | 73 (10) | 74 (10) |

^1^*ABC = Aberrant Behaviour Checklist-Community version,* ^2^*PAS-ADD = Psychiatric Assessment Schedule for Adults with Developmental Disabilities,* ^3^*TAG = Threshold Assessment Grid,* ^4^*QOL-Q = Quality of Life*

**Supplementary Table 2** Unit cost for IST models 2020/2021 (£ per year)

| Costs | Enhanced model | Independent model |
| --- | --- | --- |
| Salary | £23401 | £27477 |
| Salary oncosts including employers’ national insurance and superannuation contributions | £8050 | £9452 |
| Overheads: Management and other non-care staff | £5733 | £6732 |
| Overheads: Non-staff | £8939 | £10496 |
| Capital overheads | £5353 | £5353 |
| Ratio of direct face-to-face to indirect time | 1:0.58 | 1:0.56 |
| Working time | 33 weeks per annum and 30 hrs per week | 35 weeks per annum and 32 hrs per week |
| Caseload per team^a^ | 985 | 511 |
| Team members (mean) | 12 | 11 |
|  | £51.57 per hour; £81.48 per hour of face-to-face patient contact; £51051 average annual cost of team member; £4980 average annual cost per case. | £52.58 per hour; £82.02 per hour of face-to-face patient contact; £58892 annual average cost of team member; £10122 average annual cost per case. |

^a^ *proxy caseload calculated from referral data over 12 months represents total caseload across 8 teams.*

**Supplementary Table 3** Service use and support at baseline and 9 months follow up (for previous 6 months)

|  | | Independent Model | | | | | | | Enhanced Provision Model | | | | | | |
| --- | --- | --- | --- | --- | --- | --- | --- | --- | --- | --- | --- | --- | --- | --- | --- |
| Service use | | **Valid n** | | **Mean (bed days)** | | **(SD)** | | **% using at least once** | **Valid n** | | **Mean** | | **(SD)** | | **% using at least once** |
| Baseline | | | | | | | | | | | | | | | |
| Hospital-based care | | | | | | | | | | | | | | | |
| Psychiatric inpatient (bed day) | | 109 | | 6.94 | | 29.00 | | 10.1 | 114 | | 4.04 | | 20.20 | | 7.0 |
| General inpatient (bed day) | | 109 | | 0.27 | | 1.25 | | 7.3 | 114 | | 0.08 | | 0.75 | | 1.8 |
| Psychiatric outpatient (attendance) | | 109 | | 0.07 | | 0.35 | | 4.6 | 114 | | 0.15 | | 0.57 | | 8.8 |
| General outpatient (attendance) | | 109 | | 0.41 | | 1.23 | | 16.5 | 114 | | 0.78 | | 2.19 | | 33.1 |
| Day hospital (attendance) | | 109 | | 0.20 | | 0.62 | | 11.9 | 114 | | 0.23 | | 1.23 | | 7.0 |
| A& E Psychiatric - admitted (attendance) | | 109 | | 0.04 | | 0.19 | | 3.7 | 114 | | 0.03 | | 016 | | 2.6 |
| A& E Psychiatric - not admitted (attendance) | | 109 | | 0.14 | | 1.16 | | 3.7 | 114 | | 0.07 | | 0.37 | | 5.3 |
| A& E Physical - admitted (attendance) | | 109 | | 0.17 | | 0.99 | | 9.2 | 114 | | 0.08 | | 0.50 | | 2.6 |
| A& E Physical - not admitted (attendance) | | 109 | | 0.13 | | 0.46 | | 10.1 | 114 | | 0.47 | | 1.98 | | 20.2 |
| Community-based health and social care services | | | | | | | | | | | | | | | |
| General Practice (contacts) | | 109 | | 4.30 | | 5.90 | | 97.2 | 114 | | 4.63 | | 7.16 | | 86.8 |
| Other health and social care services (contacts) ^1^ | | 103 | | 82.9 | | 82.9 | | 100 | 108 | | 71.0 | | 100.3 | | 98.2 |
| Informal support | | | | | | | | | | | | | | | |
| Paid carer (%) | | 71 | |  | | 28 | | 39.44 | 81 | |  | | 30 | | 37 |
| 9-months follow up | | | | | | | | | | | | | | | |
| Hospital-based care | | | | | | | | | | | | | | | |
| Psychiatric inpatient (bed day) | 100 | | 8.63 | | 39.98 | | 6.0 | | 104 | 5.26 | | 28.05 | | 4.8 | |
| General inpatient (bed day) | 100 | | 0.35 | | 2.08 | | 4.0 | | 104 | 0.22 | | 1.17 | | 5.8 | |
| Psychiatric outpatient (attendance) | 100 | | 0.08 | | 0.44 | | 4.0 | | 104 | 0.23 | | 0.99 | | 8.7 | |
| General outpatient (attendance) | 100 | | 0.26 | | 0.69 | | 16.0 | | 104 | 0.40 | | 0.94 | | 22.1 | |
| Day hospital (attendance) | 100 | | 0.07 | | 0.36 | | 5.0 | | 104 | 0.03 | | 0.17 | | 2.9 | |
| A& E Psychiatric - admitted (attendance) | 100 | | 0.03 | | 0.17 | | 3.0 | | 104 | 0.02 | | 0.14 | | 1.9 | |
| A& E Psychiatric - not admitted (attendance) | 100 | | 0.01 | | 0.10 | | 1.0 | | 104 | 0.04 | | 0.24 | | 2.9 | |
| A& E Physical - admitted (attendance) | 100 | | 0.04 | | 0.24 | | 3.0 | | 104 | 0.07 | | 0.42 | | 3.9 | |
| A& E Physical - not admitted (attendance) | 100 | | 0.03 | | 0.17 | | 3.0 | | 104 | 0.07 | | 0.32 | | 4.8 | |
| Community-based health and social care services | | | | | | | | | | | | | | | |
| General Practice (contacts) | 100 | | 4.8 | | 14.20 | | 88.0 | | 104 | 3.47 | | 3.96 | | 87.5 | |
| Other health and social care services (contacts)^1^ | 100 | | 71.0 | | 100.40 | | 95.0 | | 102 | 82.19 | | 109.80 | | 96.1 | |
| Informal care | | | | | | | | | | | | | | | |
| Paid carer (yes) | 60 | |  | | 22 | | 36.67 | | 73 |  | | 30 | | 41.10 | |

**Supplementary Table 4** Mean total costs and outcomes over 9 months by IST model (£, 2020/21 prices)

|  | Independent Model (n=111) | | | Enhanced Model (n=115) | | | Independent Model – Enhanced Model | |
| --- | --- | --- | --- | --- | --- | --- | --- | --- |
| Costs | **Valid n** | **Mean** | **(SD)** | **Valid n** | **Mean** | **(SD)** | **Adjusted mean difference** | **95% CI** |
| a. IST model cost | 110 | 7591.5 | - | 115 | 3735 | - |  |  |
| b. Health and social care costs | 98 | 15324.18 | (30301.5) | 102 | 15302.66 | (25786.9) | -3409.95 | (-9957.92 to 4039.89) |
| c. Unpaid carer costs | 100 | 8963.4 | (8010) | 106 | 10520.8 | (7450.8) | -1123.01 | (-2727.50 to 621.51) |
| Total health and social care cost excluding informal care (a+b) | 98 | 22915.6 | (30301.5) | 102 | 19037.6 | (25786.9) | 446.55 | (-5637.60 to 7519.30) |
| Total health and social care cost including informal care (a+b+c) | 98 | 31850.8 | (31385.1) | 102 | 29852.8 | (26754.9) | -855.80 | (-8342.54 to 6059.69) |
| Challenging behaviour score (ABC) 9 months | 101 | 49 | (32) | 107 | 56 | (34) | -5.07 | (-14.93 to 2.14) |
| QALYs (EQ5D-proxy) 9 months | 98 | 0.5049 | (0.1574) | 101 | 0.4637 | (0.1907) | 0.0158 | (-0.0088 to 0.0508) |

1. *Adjusted mean difference on baseline total and component cost obtained following adjustment for, baseline factors such as ABC score, age, sex, accommodation type, having autism and/or ADHD, number of physical conditions. For example, comparisons of community-based costs at baseline for the models include adjustment for baseline ABC score, age, sex, accommodation type, having autism and/or ADHD, number of physical conditions.*

2. *Adjusted mean difference on total and component costs at 9 months obtained following adjustment for baseline cost measure and baseline factors such as ABC score, age, sex, accommodation type, having autism and/or ADHD, number of physical conditions. For example, comparisons of community-based costs at 9 months for the models include adjustment for, baseline community-based costs, ABC score, baseline age, sex, accommodation type, having autism and/or ADHD, number of physical conditions*

**Supplementary Table 5** Differences in incremental costs, effect and cost-effectiveness

|  | Health and social care perspective | Societal perspective |
| --- | --- | --- |
| Incremental costs £, mean (95% CI) ^a^ | 446.55 (-5637.60 to 7519.30) | -855.80 (-8342.54 to 6059.69) |
| Incremental effect mean (95% CI): | | |
| Points Improvement in ABC score | -5.07 (-14.93 to 2.14) | -5.07 (-14.93 to 2.14) |
| QALY EQ-5D, proxy score | 0.0158 (-0.0088 to 0.0508) | 0.0158 (-0.0088 to 0.0508) |
| ICER: | | |
| Additional cost per additional point improvement in the ABC scale (£) | 88.08 | -168.79 |
| Additional cost per additional QALY (£) | 28262.66 | -54164.56 |

^a^ *Includes cost of the IST model*

#### Figures

**Supplementary Figure 1** Cost-effectiveness acceptability curve showing the probability that Independent model is cost-effective compared with Enhanced; health and social care perspective, with effectiveness measured in ABC score at 9 months

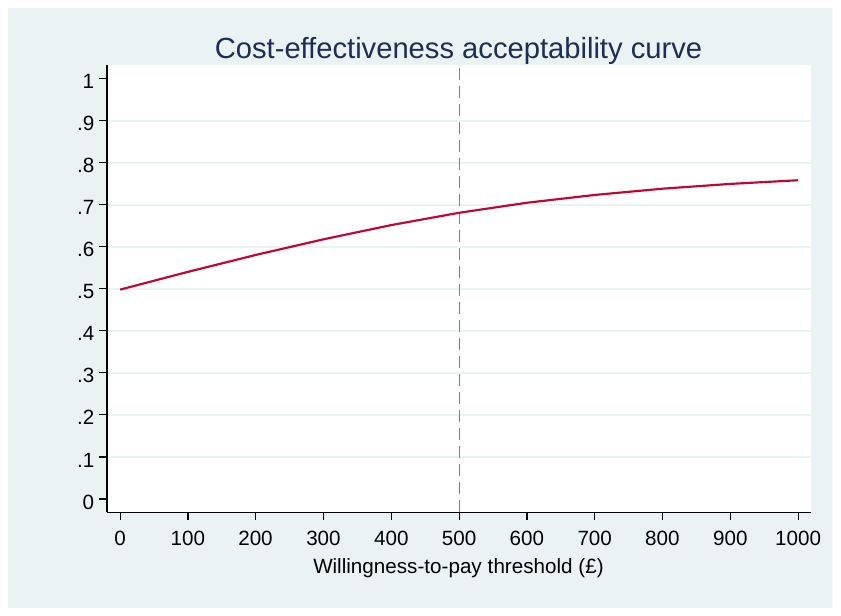

**Supplementary Figure 2** Cost-effectiveness acceptability curve showing the probability that independent model is cost-effective compared with enhanced model; societal perspective, with effectiveness measured in ABC-C score at 9 months and societal costs adjusted

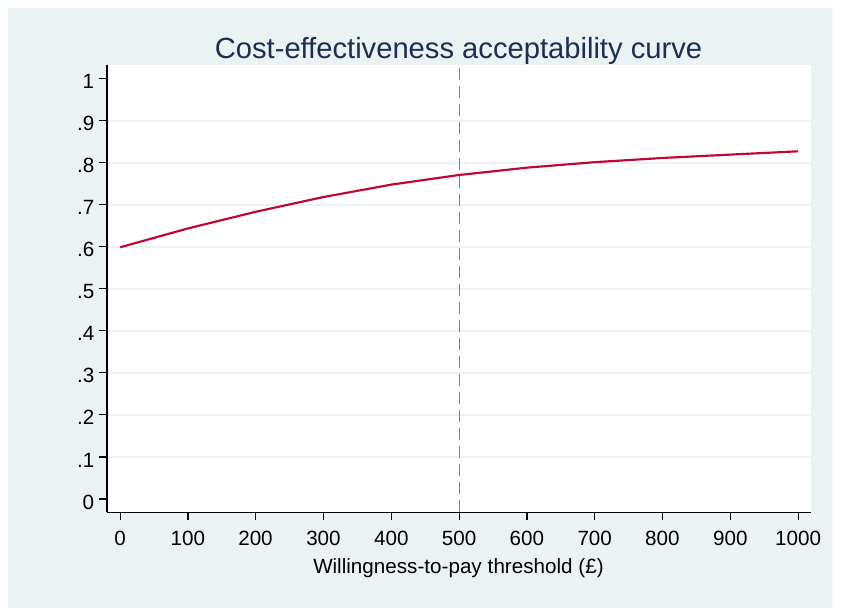

**Supplementary Figure 3** Cost-effectiveness acceptability curve showing the probability that Independent model is cost-effective compared with Enhanced model; health and social care perspective, with effectiveness measured in QALYs over 9 months

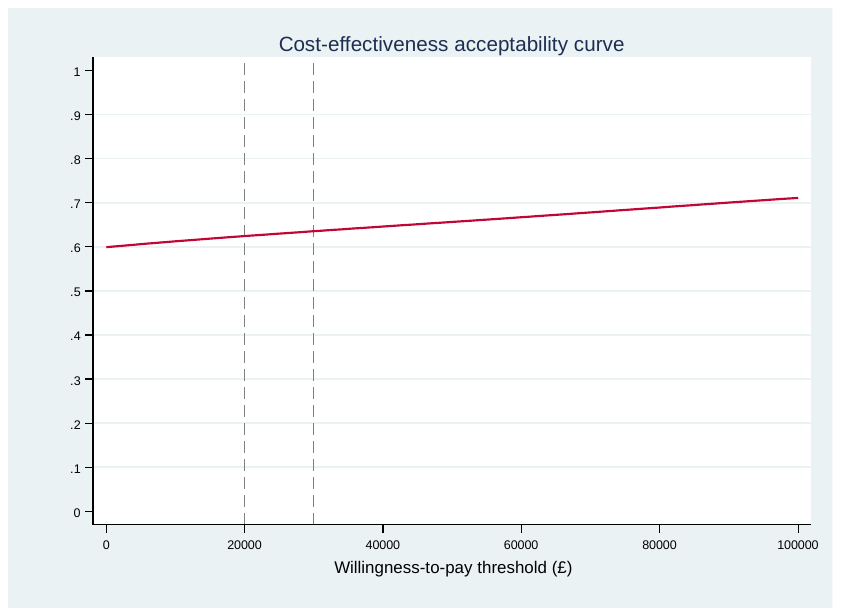

**Supplementary Figure 4** Cost-effectiveness acceptability curve showing the probability that Independent model is cost-effective compared with Enhanced; societal perspective, with effectiveness measured in QALYS over 9 months and societal costs adjusted

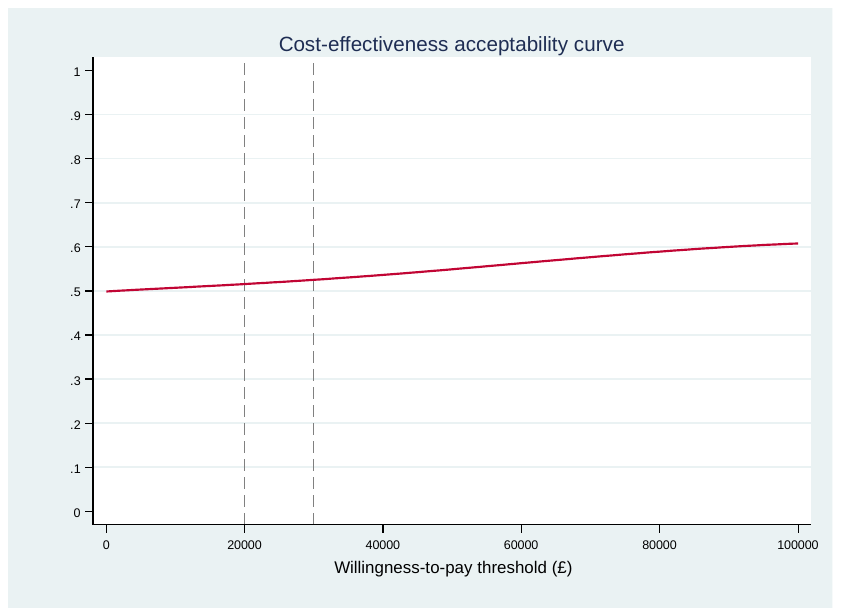
